# Correlates of Ki-67 Proliferation Index in a Cohort of Women with Suspected Breast Cancer in Lusaka, Zambia

**DOI:** 10.64898/2025.12.16.25342441

**Authors:** Joaquim Musamba, David Chisompola, Situmbeko Liweleya, Kingsley Kamvuma, Phinnoty Mwansa, Sepiso K. Masenga

## Abstract

**Introduction:** Ki-67 is a key biomarker of tumor proliferation in breast cancer, yet its clinical correlates in breast cancer screening populations, where disease is often detected at earlier stages, and remain underexplored. This study aimed to identify factors independently associated with high Ki-67 expression among women investigated for suspected breast cancer at Unilabs Laboratory, Lusaka, Zambia.

**Methods:** A retrospective cross-sectional analysis was conducted on 208 women suspected with breast cancer through a laboratory-based screening program in Lusaka, Zambia (2019–2024). Demographic, clinical, and pathological data were extracted from laboratory records, and Ki-67 expression was dichotomized as low (<20%) or high (>20%). Variables significant in Bivariate analysis (p<0.05) were included in a multivariable logistic regression model to identify independent predictors of high Ki-67 expression. Variables significant in bivariate analysis (p<0.05) were included in multivariable logistic regression models to identify factors associated with high Ki-67 expression.

**Results:** The median age was 48 years (IQR: 40–63), and 46.2% (n=96) exhibited high Ki-67 expression. In bivariate analysis, younger age (<40 years), invasive ductal carcinoma, right breast involvement, progesterone receptor (PR) positivity threshold ≥10%, non-Quick Score scoring methods, and lack of fluorescence in situ hybridization (FISH) testing were associated with high Ki-67. However, in adjusted multivariable models, only younger age (>41 years) (aOR: 0.43–0.46, p<0.05), PR positivity threshold ≥10% (aOR: 6.14–6.38, p<0.05), and use of non-Quick Score scoring methods (aOR: 10.5–14.9, p≤0.002) remained significantly associated with high Ki-67, while other factors lost statistical significance after controlling for confounders.

**Conclusion:** In this diagnostic cohort from Zambia, nearly half of women with breast cancer exhibited high Ki-67 expression, reinforcing its relevance even in early detection settings. The study identified younger age, higher PR positivity threshold, and alternative scoring methods as independent predictors of high Ki-67, highlighting the importance of standardized biomarker assessment. Future studies should consider prospective study design incorporating molecular subtyping to enhance the clinical interpretation of Ki-67 in similar populations.

## Introduction

Breast cancer remains the most frequently diagnosed cancer among women worldwide, with an estimated 685,000 deaths reported in 2020 [1–4]. The global burden of breast cancer continues to rise, driven by aging populations, lifestyle changes, and improvements in cancer detection and reporting [5,6]. Robust screening programs are therefore critical for early diagnosis and improved survival outcomes.

The management of breast cancer has been increasingly guided by personalized medicine, relying on biomarker-driven stratification [7]. Established biomarkers, including estrogen receptor (ER), progesterone receptor (PR), and human epidermal growth factor receptor 2 (HER2), are essential for prognostication and for tailoring targeted therapies, thereby enhancing treatment efficacy and minimizing unnecessary toxicity [2,8–11]. Beyond these, the Ki-67 proliferation index has emerged as an important supplementary biomarker [7,12]. As a nuclear protein expressed during all active phases of the cell cycle, Ki-67 reflects the proportion of proliferating tumor cells, providing a direct measure of tumor aggressiveness. Elevated Ki-67 expression has been consistently linked with more aggressive tumor biology, poorer prognosis, and greater responsiveness to certain chemotherapeutic regimens [13,14].

Despite its clinical relevance, the routine application of Ki-67 faces challenges, particularly regarding standardization and interpretation [7,15]. Most evidence on Ki-67’s prognostic value derives from studies of cohorts with confirmed, often advanced, breast cancer [16–19]. There is limited evidence from sub-Saharan Africa, and in Zambia on Ki-67 expression patterns in screening populations, where disease stage and biomarker behavior may differ from clinical cohorts. This lack of local, screening-population data hinders the effective integration of Ki-67 into routine prognostic models and treatment decision-making in our setting. Understanding these associations is essential to optimize Ki-67’s utility across the full spectrum of breast disease.

This gap in knowledge presents a challenge for the standardized application of Ki-67 in clinical practice. A more comprehensive understanding of its correlations in a heterogeneous screening population could improve interpretation and contextualization of this biomarker. Therefore, this study aimed to systematically characterize the clinical and pathological factors associated with Ki-67 expression in a cohort of women with clinically suspected breast cancer who underwent diagnostic biopsy at Unilabs, Lusaka, Zambia.

## Methodology

### Design, Setting and Population

This retrospective cross-sectional study utilized laboratory data from women with suspected breast cancer whose diagnostic samples were processed at Unilabs Laboratory in Lusaka, Zambia, between January 2019 and December 2024. Data were abstracted from electronic medical records between 02 June and 31 July, 2025. A total of 210 records were reviewed, of which 208 met the inclusion criteria and were enrolled in the study; two (2) were excluded as they were male participants.

### Inclusion and exclusion criteria

Eligibility criteria were defined to ensure consistency and reliability of the study data. The study included histologically confirmed cases of breast cancer among women with documented expression levels of key biomarkers, namely estrogen receptor (ER), progesterone receptor (PR), human epidermal growth factor receptor 2 (HER2), and the proliferation marker Ki-67. Cases were excluded if biomarker expression data were incomplete or missing. Additionally, patients who had received neoadjuvant chemotherapy or hormone therapy prior to biomarker assessment were excluded to avoid potential alterations in baseline expression levels. Male patients were also excluded from the study.

### Study Variables

The variables collected encompassed demographic, clinical, and pathological characteristics. Demographic variables included age and menopausal status. Clinical presentation data captured self-reported or clinically documented information regarding the affected breast side. Pathological variables included final diagnosis (cancer versus non-cancer), histological subtype, and tumor grade. Biomarker-related variables consisted of estrogen receptor (ER), progesterone receptor (PR), and human epidermal growth factor receptor 2 (HER2) status, including staining intensity and the positivity threshold. However, personal identifiers such as names were not retrieved from the electronic registers in order to ensure anonymity of the participants.

Ki-67 expression was assessed immunohistochemically on core biopsy or surgical specimens, and patients were categorized into high or low Ki-67 expression groups based on a predefined institutional cutoff (0: Low Proliferation <20%, 1: High Proliferation >20%) [20]. Additional information regarding the biomarker scoring method (such as Quick Score or alternative systems) and the performance of fluorescence in situ hybridization (FISH) testing for equivocal HER2 results was also recorded.

### Data Collection

Clinical and pathological data were retrospectively extracted from electronic medical records from June 2nd to July 31st, 2025, and systematically entered into a standardized electronic report form using the REDCap data management platform. Following extraction, the dataset was exported to Microsoft Excel 2019 for data cleaning and validation prior to statistical analysis.

### Statistical Analysis

Data are presented as medians with interquartile ranges (IQR) for continuous variables and frequencies with percentages for categorical variables. The Mann–Whitney U test was used for continuous variables, and the Chi-square or Fisher’s exact test was used for categorical variables to assess bivariate associations with Ki-67 expression status (dichotomized as low <20% or high 20%). Variables with a p-value <0.05 in bivariate analysis were entered into multivariable logistic regression models to identify factors independently associated with high Ki-67 expression. To account for potential confounding and to assess the robustness of the associations, three separate multivariable models were fitted:

- Model 1 adjusted for estrogen receptor (ER) status;
- Model 2 adjusted for progesterone receptor (PR) status;
- Model 3 adjusted for human epidermal growth factor receptor 2 (HER2) status.

Each model included the same core set of covariates: age group (>41 years vs. ≤40 years), histology type (invasive ductal carcinoma vs. non-carcinoma), clinical history (affected side) (right vs. left breast), PR positivity threshold (≥10% vs. >1%), biomarker scoring method (Quick Score vs. other), and fluorescence in situ hybridization (FISH) testing status (yes vs. no). All analyses were performed using Stata version 15, and a p-value < 0.05 was considered statistically significant.

### Ethical consideration and consent to participate

Ethical clearance for this study was granted by the Mulungushi University School of Medicine and Health Sciences (SOMHS) Research Ethics Committee (Reference No: SMHS-MU2-2025-14) on 6^th^ May 2025, and by the National Health Research Authority (Reference No. NHRA-2208/08/05/2025). The study utilized secondary data extracted from existing medical records; therefore, no personally identifiable information was collected or included in the data collection forms. As the study involved retrospective analysis of anonymized data, the requirement for written or verbal informed consent was waived by the ethics committees.

The study was conducted in accordance with the principles of the Declaration of Helsinki and the Good Clinical Practice (GCP) Guidelines. Reporting adhered to the Strengthening the Reporting of Observational Studies in Epidemiology (STROBE) checklist, as detailed in Supplementary File S1. Additionally, the anonymized dataset used for the analysis is provided as supporting information in File S2.

## Results

### Patient and Tumor Characteristics

A total of 208 women were included in this study. The cohort’s demographic, clinical, and pathological characteristics are summarized in Table 1. The median age of the participants was 48 years (IQR: 40, 63). The majority of patients were postmenopausal (62.5%, n=130) and had a diagnosis of breast cancer (94.2%, n=196), with invasive ductal carcinoma (IDC) being the predominant histology (94.2%, n=195). Based on the study’s Ki-67 cutoff, 96 patients (46.2%) were classified as having high Ki-67 expression, while 112 (53.8%) had low expression Table 1.

**Table 1.** Association Between Clinical/Pathological Factors and Ki-67 Expression in Women.

| Characteristic | Median (IQR) /Frequency (%) | Ki-67 Expression |  | p-value |
| --- | --- | --- | --- | --- |
|  |  | High = 96 (46.2 %) | Low = 112 (53.8%) |  |
| **Demographics** |  |  |  |  |
| **Age (years)** | 48 (40, 63) | 46 (38,62.7) | 50.6 (42, 63) | 0.0767 |
| **Age Group (years)** |  |  |  |  |
| *<40 years* | 57 (27.4) | 34 (59.6) | 23 (40.4) | **0.016** |
| *> 41 years* | 151 (72.6) | 62 (41.1) | 89 (58.9) |  |
| **Menopausal Status** |  |  |  |  |
| *Premenopausal* | 78 (37.5) | 41 (52.6) | 37 (47.4) | 0.151 |
| *Postmenopausal* | 130 (62.5) | 55 (42.3) | 75 (57.7) |  |
| **Histology Type** |  |  |  |  |
| *IDC* | 195 (94.2) | 83 (42.6) | 112 (57.4) | **<0.0001** |
| *No Carcinoma* | 5 (2.4) | 5 (100) | 0 (0) |  |
| **Clinical Presentation** |  |  |  |  |
| **Clinical History (Affected Side)** |  |  |  |  |
| *Left Breast* | 29 (13.9) | 6 (20.7) | 23 (79.3) | **0.007** |
| *Right Breast* | 35 (16.8) | 15 (42.9) | 20 (57.1) |  |
| **Pathology** |  |  |  |  |
| **Tumor Grade** |  |  |  |  |
| *Grade 2* | 1 (0.5) | 1 (100) | 0 (0) | 0.170 |
| *Grade 3* | 11 (5.3) | 3 (27.3) | 8 (72.7) |  |
| **Biomarker Status** |  |  |  |  |
| **ER Status** |  |  |  |  |
| *Negative* | 84 (40.4) | 34 (40.5) | 50 (59.5) | 0.176 |
| *Positive* | 124 (59.6) | 62 (50.0) | 62 (50.0) |  |
| **PR Status** |  |  |  |  |
| *Negative* | 117 (56.3) | 52 (44.4) | 65 (55.6) | 0.575 |
| *Positive* | 91 (43.7) | 44 (48.4) | 47 (51.6) |  |
| **HER2 Status** |  |  |  |  |
| *Negative* | 132 (63.5) | 62 (47.0) | 70 (53.0) | 0.756 |
| *Positive* | 76 (36.5) | 34 (44.7) | 42 (55.3) |  |
| **Biomarker Detailed Staining** |  |  |  |  |
| **ER Staining Intensity** |  |  |  |  |
| *Strong* | 93 (44.7) | 46 (49.5) | 47 (50.5) | 0.647 |
| *Moderate* | 9 (4.3) | 5 (55.6) | 4 (44.4) |  |
| *Weak* | 23 (11.1) | 11 (47.8) | 12 (52.2) |  |
| **ER Positivity Threshold** |  |  |  |  |
| >1% | 19 (9.1) | 6 (31.6) | 13 (68.4) | 0.189 |
| >10% | 37 (17.8) | 21 (56.8) | 16 (43.2) |  |
| **PR Staining Intensity** |  |  |  |  |
| *Strong* | 46 (22.1) | 26 (56.5) | 20 (43.5) | 0.430 |
| *Moderate* | 8 (3.9) | 3 (37.5) | 5 (62.5) |  |
| *Weak* | 32 (15.4) | 13 (40.6) | 19 (59.4) |  |
| **PR Positivity Threshold** |  |  |  |  |
| >1% | 18 (8.6) | 4 (22.2) | 14 (77.8) | **0.038** |
| >10% | 17 (8.2) | 11 (64.7) | 6 (35.3) |  |
| **Scoring Method** |  |  |  |  |
| *Quick Score* | 192 (92.3) | 82 (42.7) | 110 (57.3) | **0.001** |
| *Other* | 16 (7.7) | 14 (87.5) | 2 (12.5) |  |
| **HER2 Positivity Threshold** |  |  |  |  |
| >10% | 21 (10.1) | 8 (38.1) | 13 (61.9) | 0.626 |
| >30% | 15 (7.2) | 6 (40.0) | 9 (60.0) |  |
| **FISH Testing** |  |  |  |  |
| *Yes* | 4 (1.9) | 1 (25.0) | 3 (75.0) | **<0.0001** |
| *No* | 190 (91.4) | 82 (43.2) | 108 (56.8) |  |
| **Abbreviations:** Median (IQR; Interquartile Range) or Frequency (%; Percentage). P-values for continuous variables (Age) were calculated using the Mann-Whitney U test; p-values for categorical variables were calculated using the Chi-square or Fisher's exact test, as appropriate. P-values in Bold means it is statistically significant (<0.05)<br>a. ER: Estrogen Receptor; PR: Progesterone Receptor; IDC: Invasive ductal carcinoma<br>b. The "No Cancer" group includes benign breast conditions. The "No Carcinoma" and "Other" histology groups include non-malignant diagnoses and non-IDC malignancies, respectively. |  |  |  |  |

### Factors Associated with High Ki-67 Expression

Univariable and multivariable logistic regression analyses were performed to identify clinicopathological factors associated with high Ki-67 expression. Three separate multivariable models (Models 1, 2 and 3) were constructed, each incorporating a different primary biomarker (ER status, PR status, and HER2 status, respectively) while adjusting for the same core set of variables: age, histology type, tumor laterality, PR positivity threshold, scoring method, and FISH testing status. The results are presented in Tables 2. In univariable analysis, which was consistent across all models, several factors were significantly associated with high Ki-67 status. Patient age greater than 41 years was inversely associated with high Ki-67 (odds ratio [OR]: 0.47, 95% confidence interval [CI]: 0.25–0.87, p=0.017). Positive associations were observed for a progesterone receptor (PR) positivity threshold >10% (OR: 6.41, 95% CI: 1.44–28.5, p=0.015) and the use of a scoring method other than “Quick Score” (OR: 10.1, 95% CI: 2.26–45.7, p=0.002) (Tables 2). A non-significant trend towards higher Ki-67 was noted for tumors in the right breast (OR: 2.87, 95% CI: 0.93–8.81, p=0.065). For the histology category “No Carcinoma,” quasi-complete separation of data points occurred, preventing valid odds ratio calculation. The multivariable analyses confirmed the independence of three key predictors, with remarkably consistent effect sizes across all three models (Tables 2). Age >41 years remained a significant protective factor (adjusted OR [aOR] range across models: 0.43 to 0.46; all p < 0.05). Conversely, PR positivity threshold >10% (aOR range: 6.14 to 6.38; all p < 0.05) and use of an alternative scoring method (aOR range: 10.5 to 14.9; all p ≤ 0.002) were strong, independent predictors of high Ki-67 expression. The association for right breast tumors was attenuated and remained non-significant in the multivariable models (aOR range: 2.87 to 3.09; p=0.050 to 0.065). FISH testing status was not a significant predictor in any model. The issue of quasi-complete separation for the “No Carcinoma” group persisted in the multivariable analyses (Tables 2).

**Table 2.** Univariable and Multivariable Logistic Regression Analyses of Factors Associated with High Ki-67 Expression.

| Variable | OR (95% CI) | p-value | Model 1 (ER)<br>aOR (95% CI) | p-value | Model 2 (PR)<br>aOR (95% CI) | p-value | Model 3 (HER2)<br>aOR (95% CI) | p-value |
| --- | --- | --- | --- | --- | --- | --- | --- | --- |
| **Age Group** |  |  |  |  |  |  |  |  |
| *<40 years* | Ref | – | Ref | – | Ref | – | Ref | – |
| *>41 years* | 0.47 (0.25, 0.87) | **0.017** | 0.46 (0.24, 0.86) | **0.016** | 0.45 (0.24, 0.85) | **0.014** | 0.43 (0.23, 0.83) | **0.012** |
| **Histology** |  |  |  |  |  |  |  |  |
| IDC | Ref | – | Ref | – | Ref | – | Ref | – |
| No Carcinoma | † | – | † | – | † | – | † | – |
| **Clinical History**<br>**(Side affected)** |  |  |  |  |  |  |  |  |
| *Left Breast* | Ref | – | Ref | – | Ref | – | Ref | – |
| *Right Breast* | 2.87 (0.93, 8.81) | 0.065 | 3.09 (0.99, 9.57) | 0.050 | 2.89 (0.94, 8.90) | 0.063 | 2.87 (0.93, 8.81) | 0.065 |
| **PR Threshold** |  |  |  |  |  |  |  |  |
| *>1%* | Ref | – | Ref | – | Ref | – | Ref | – |
| *>10%* | 6.41 (1.44, 28.5) | **0.015** | 6.14 (1.37, 27.3) | **0.017** | 6.16 (1.37, 27.6) | **0.017** | 6.38 (1.43, 28.5) | **0.015** |
| **Scoring Method** |  |  |  |  |  |  |  |  |
| *Quick Score* | Ref | – | Ref | – | Ref | – | Ref | – |
| *Other* | 10.1 (2.26, 45.7) | 0.002 | 14.9 (3.18–70.2) | 0.001 | 12.2 (2.65–56.1) | 0.001 | 10.5 (2.32–48.2) | 0.002 |
| **FISH Testing** |  |  |  |  |  |  |  |  |
| *Yes* | Ref | – | Ref | – | Ref | – | Ref | – |
| *No* | 2.27 (0.23, 22.2) | 0.479 | 3.10 (0.31, 30.6) | 0.333 | 2.23 (0.23–23.0) | 0.470 | 2.41 (0.24–23.8) | 0.451 |
| Abbreviations: OR, odds ratio; aOR, adjusted odds ratio; CI, confidence interval; ER, estrogen receptor; HER2, human epidermal growth factor receptor 2; IDC, invasive ductal carcinoma; OR, odds ratio; PR, progesterone receptor. Note: All multivariable models were adjusted for all variables listed in the table, with the primary biomarker (ER, PR, or HER2 status) integrated as appropriate for each model. † Odds ratio not calculable due to quasi-complete separation |  |  |  |  |  |  |  |  |

## Discussion

This study aimed to identify factors associated with a high Ki-67 proliferation index among women investigated for suspected breast cancer in Lusaka, Zambia. The analysis revealed several significant associations in univariable models, and three factors remained independently associated with high Ki-67 in adjusted multivariable analyses: younger age (>41 years), a progesterone receptor (PR) positivity threshold of ≥10%, and the use of a biomarker scoring method other than the Quick Score. These findings provide new insights into the clinicopathological correlates of Ki-67 expression in a diagnostic cohort from sub-Saharan Africa and underscore both the relevance and the complexity of interpreting this biomarker in clinical practice.

Our study reveals a 46.2% prevalence of high Ki-67 expression in this diagnostic cohort of women with breast cancer. This proportion is lower than the 56.5% and 50.5% reported in the monarchE Phase 3 clinical trial [21,22]. This variation may reflect differences in tumor biology, population, or the Ki-67 scoring thresholds applied. In contrast, it is higher than the 33.8% reported in a study from Pakistan [23], suggesting potential geographic, genetic, or environmental influences on tumor proliferative activity. Overall, nearly half of the screened women in our setting exhibited high-proliferation tumors, reinforcing the importance of Ki-67 as a prognostic biomarker even in early-detection contexts.

In the adjusted multivariable models, age >41 years was inversely associated with high Ki-67 (aOR range: 0.43–0.46, p < 0.05). This aligns with evidence that younger women often present with more aggressive, rapidly proliferating tumors [24]. The strong independent association between a PR positivity threshold of ≥10% and high Ki-67 (aOR range: 6.14–6.38, p < 0.05) is noteworthy, as PR expression is typically linked to less aggressive, hormone-responsive disease [25]. This counter-intuitive finding may reflect unique tumor biology in our population or indicate that high PR expression, when coupled with high proliferation, identifies a distinct subgroup with altered hormonal signaling.

The study findings reveals the use of a scoring method other than the Quick Score as a predictor of high ki-67 expression (aOR range: 10.5–14.9, p ≤ 0.002). This suggests a methodological issue where variability in biomarker assessment can profoundly influence Ki-67 categorization. These issues include inconsistencies in scoring practices, whether due to different interpretation guidelines, laboratory protocols, or pathologist experience, which may introduce significant bias and reduce the comparability of Ki-67 results across studies and clinical settings. Our finding underscores the important need for standardized, validated scoring systems to ensure reliable and reproducible Ki-67 reporting.

Clinically, high Ki-67 expression is a well-established marker of tumor aggressiveness, correlating with poorer prognosis, higher histological grade, and increased risk of recurrence [27–30]. Our results affirm that Ki-67 remains a relevant biomarker in a Zambian screening population. However, the strong dependence on scoring methodology observed here calls for caution in its clinical application. Without local standardization, Ki-67 results may be misleading and could potentially affect treatment decisions, particularly in resource-limited settings where access to advanced molecular subtyping is limited.

### Strengths and Limitations

This study has several strengths, including a well-characterized cohort, rigorous multivariable modeling adjusted for key biomarkers, and transparent reporting of both univariable and multivariable associations. However, important limitations must be acknowledged. The cross-sectional design precludes causal inference. Quasi-complete separation for the “No Carcinoma” category limited the inclusion of this group in regression models. The absence of molecular subtyping data restricts our ability to contextualize Ki-67 within established intrinsic subtypes. Additionally, the retrospective reliance on laboratory records may have introduced variability in biomarker assessment and documentation. Future prospective studies, incorporating standardized scoring protocols and molecular classification, are needed to validate these findings and better define the role of Ki-67 in similar populations.

## Conclusion

In this Zambian diagnostic cohort, nearly half of the women exhibited high Ki-67 expression. Younger age, a higher PR positivity threshold, and the use of non-Quick Score scoring methods were independent predictors of high Ki-67. These findings underscore both the clinical relevance of Ki-67 and the critical importance of standardized biomarker assessment in ensuring its reliable interpretation. Efforts to harmonize scoring practices and integrate molecular subtyping in future studies will enhance the utility of Ki-67 in prognostic stratification and treatment planning in sub-Saharan African settings.

## Availability of data and materials

The raw data underlying the results presented in the study have been uploaded as Supporting information.

## Competing interests

The authors have declared that no competing interests exist

## Funding

This study received no funding

## Supplementary files

S1. Strobe checklist

S2. Data

## Notes

### Competing Interest Statement

The authors have declared no competing interest.

### Author Declarations

Ethical clearance for this study was granted by the Mulungushi University School of Medicine and Health Sciences (SOMHS) Research Ethics Committee (Reference No: SMHS-MU2-2025-14) on 6th May 2025, and by the National Health Research Authority (Reference No. NHRA-2208/08/05/2025). The study utilized secondary data extracted from existing medical records therefore, no personally identifiable information was collected or included in the data collection forms. As the study involved retrospective analysis of anonymized data, the requirement for written or verbal informed consent was waived by the ethics committees. The study was conducted in accordance with the principles of the Declaration of Helsinki and the Good Clinical Practice (GCP) Guidelines. Reporting adhered to the Strengthening the Reporting of Observational Studies in Epidemiology (STROBE) checklist

## References

1. Cabanes A, Kapambwe S, Citonje-Msadabwe S, Parham GP, Lishimpi K, Cruz TA, et al. Challenges, Opportunities, and Priorities for Advancing Breast Cancer Control in Zambia: A Consultative Meeting on Breast Cancer Control. JGO. 2019;1–7. 10.1200/JGO.18.00222

2. Hu X, Chen W, Li F, Ren P, Wu H, Zhang C, et al. Expression changes of ER, PR, HER2, and Ki-67 in primary and metastatic breast cancer and its clinical significance. Front Oncol. 2023;13:1053125. 10.3389/fonc.2023.1053125

3. Tien TZ, Lee JNLW, Lim JCT, Chen X-Y, Thike AA, Tan PH, et al. Delineating the breast cancer immune microenvironment in the era of multiplex immunohistochemistry/immunofluorescence. Histopathology. 2021;79:139–59. 10.1111/his.14328

4. Yaghoobi V, Martinez-Morilla S, Liu Y, Charette L, Rimm DL, Harigopal M. Advances in quantitative immunohistochemistry and their contribution to breast cancer. Expert Rev Mol Diagn. 2020;20:509–22. 10.1080/14737159.2020.1743178

5. Arnold M, Morgan E, Rumgay H, Mafra A, Singh D, Laversanne M, et al. Current and future burden of breast cancer: Global statistics for 2020 and 2040. Breast. 2022;66:15–23. 10.1016/j.breast.2022.08.010

6. Liu S, Tang Y, Li J, Zhao W. Global, regional, and national trends in the burden of breast cancer among individuals aged 70 years and older from 1990 to 2021: an analysis based on the global burden of disease study 2021. Archives of Public Health. 2024;82:170. 10.1186/s13690-024-01404-3

7. Davey MG, Hynes SO, Kerin MJ, Miller N, Lowery AJ. Ki-67 as a Prognostic Biomarker in Invasive Breast Cancer. Cancers (Basel). 2021;13:4455. 10.3390/cancers13174455

8. Grabinski N, Möllmann K, Milde-Langosch K, Müller V, Schumacher U, Brandt B, et al. AKT3 regulates ErbB2, ErbB3 and estrogen receptor α expression and contributes to endocrine therapy resistance of ErbB2(+) breast tumor cells from Balb-neuT mice. Cell Signal. 2014;26:1021–9. 10.1016/j.cellsig.2014.01.018

9. Thuc Nguyen TM, Dinh Le R, Nguyen CV. Breast cancer molecular subtype and relationship with clinicopathological profiles among Vietnamese women: A retrospective study. Pathology - Research and Practice. 2023;250:154819. 10.1016/j.prp.2023.154819

10. Hammond MEH, Hayes DF, Dowsett M, Allred DC, Hagerty KL, Badve S, et al. American Society of Clinical Oncology/College of American Pathologists Guideline Recommendations for Immunohistochemical Testing of Estrogen and Progesterone Receptors in Breast Cancer. Arch Pathol Lab Med. 2010;134:907–22. 10.5858/134.6.907

11. Wolff AC, Hammond MEH, Hicks DG, Dowsett M, McShane LM, Allison KH, et al. Recommendations for Human Epidermal Growth Factor Receptor 2 Testing in Breast Cancer: American Society of Clinical Oncology/College of American Pathologists Clinical Practice Guideline Update. Arch Pathol Lab Med. 2013;138:241–56. 10.5858/arpa.2013-0953-SA

12. Sağmen SB, Doğan C, Cömert S, Kıral N, Parmaksız ET, Fidan A, et al. The importance of Ki-67 proliferation index in small cell lung cancer. The Egyptian Journal of Bronchology. 2025;19:4. 10.1186/s43168-024-00357-z

13. Jung EJ, Kim J-Y, Kim J-M, Lee HS, Kwag S-J, Park J-H, et al. Positive estrogen receptor status is a poor prognostic factor in node-negative breast cancer: An observational study in Asian patients. Medicine. 2021;100:e25000. 10.1097/MD.0000000000025000

14. Kutomi G, Mizuguchi T, Satomi F, Maeda H, Shima H, Kimura Y, et al. Current status of the prognostic molecular biomarkers in breast cancer: A systematic review. Oncology Letters. 2017;13:1491–8. 10.3892/ol.2017.5609

15. Polley M-YC, Leung SCY, Gao D, Mastropasqua MG, Zabaglo LA, Bartlett JMS, et al. An international study to increase concordance in Ki67 scoring. Mod Pathol. Nature Publishing Group; 2015;28:778–86. 10.1038/modpathol.2015.38

16. Khande T, Joshi A, Khandeparkar SS, Kulkarni M, Gogate B, Kakade A, et al. Study of ER, PR, HER2/neu, p53, and Ki67 expression in primary breast carcinomas and synchronous metastatic axillary lymph nodes. Indian J Cancer. 2020;57:190. 10.4103/ijc.IJC_610_18

17. Kobayashi T, Iwaya K, Moriya T, Yamasaki T, Tsuda H, Yamamoto J, et al. A simple immunohistochemical panel comprising 2 conventional markers, Ki67 and p53, is a powerful tool for predicting patient outcome in luminal-type breast cancer. BMC Clin Pathol. 2013;13:5. 10.1186/1472-6890-13-5

18. Yerushalmi R, Woods R, Ravdin PM, Hayes MM, Gelmon KA. Ki67 in breast cancer: prognostic and predictive potential. The Lancet Oncology. 2010;11:174–83. 10.1016/S1470-2045(09)70262-1

19. Nielsen TO, Leung SCY, Rimm DL, Dodson A, Acs B, Badve S, et al. Assessment of Ki67 in Breast Cancer: Updated Recommendations From the International Ki67 in Breast Cancer Working Group. J Natl Cancer Inst. 2021;113:808–19. 10.1093/jnci/djaa201

20. Kretzmann HG, Adeniyi OV. Clinicopathological and molecular subtypes of breast cancer in the Eastern Cape, South Africa: A two-year retrospective study. PLOS ONE. Public Library of Science; 2025;20:e0325387. 10.1371/journal.pone.0325387

21. Polewski MD, Nielsen GB, Gu Y, Weaver AT, Gegg G, Tabuena-Frolli S, et al. A Standardized Investigational Ki-67 Immunohistochemistry Assay Used to Assess High-Risk Early Breast Cancer Patients in the monarchE Phase 3 Clinical Study Identifies a Population With Greater Risk of Disease Recurrence When Treated With Endocrine Therapy Alone. Appl Immunohistochem Mol Morphol. 2022;30:237–45. 10.1097/PAI.0000000000001009

22. Harbeck N, Johnston S, Fasching P, Martin M, Toi M, Rastogi P, et al. Abstract PD2-01: High Ki-67 as a biomarker for identifying patients with high risk early breast cancer treated in monarchE. Cancer Res. 2021;81:PD2–01. 10.1158/1538-7445.SABCS20-PD2-01

23. Wajid S, Samad FA, Syed AS, Kazi F, Wajid S, Samad FA, et al. Ki-67 and Its Relation With Complete Pathological Response in Patients With Breast Cancer. Cureus [Internet]. Cureus; 2021 [cited 2025 Oct 4];13. 10.7759/cureus.16788

24. Henzler M, Willborn KC, Janni W, Huober J, Lukac S, Otremba B, et al. Oncologic Outcomes of Young Breast Cancer Patients According to Tumor Biology. Cancers. 2025;17:1333. 10.3390/cancers17081333

25. Lu Z, Yang J, Feng Y, Ming J. Integrated proteomics and transcriptomics analysis reveals key regulatory genes between ER-positive/PR-positive and ER-positive/PR-negative breast cancer. BMC Cancer. 2025;25:1048. 10.1186/s12885-025-14451-y

26. Okecha T, Yahaya JJ, Waiswa A, Nyakato V, Mawanda A, Bogere N, et al. Ki67 Expression and Its Association With Clinicopathological Factors and Intrinsic Subtypes of Male Breast Cancer in Uganda: A Cross-sectional Study. Breast Cancer (Auckl). 2025;19:11782234251392693. 10.1177/11782234251392693

27. Muftah AA, Aleskandarany MA, Al-kaabi MM, Sonbul SN, Diez-Rodriguez M, Nolan CC, et al. Ki67 expression in invasive breast cancer: the use of tissue microarrays compared with whole tissue sections. Breast Cancer Res Treat. 2017;164:341–8. 10.1007/s10549-017-4270-0

28. Sandilya U. K M. Expression of Ki-67 in Invasive Breast Carcinoma and Its Correlation With Different Clinicopathological Features. Cureus. 16:e69820. 10.7759/cureus.69820

29. Wongmaneerung P, Chitapanarux I, Traisathit P, Prasitwattanaseree S, Rottuntikarn W, Somwangprasert A, et al. The association between Ki-67 expression and survival in breast cancer subtypes: a cross-sectional study of Ki-67 cut-point in northern Thailand. BMC Cancer. 2025;25:346. 10.1186/s12885-025-13724-w

30. Liang Q, Ma D, Gao R-F, Yu K-D. Effect of Ki-67 Expression Levels and Histological Grade on Breast Cancer Early Relapse in Patients with Different Immunohistochemical-based Subtypes. Sci Rep. Nature Publishing Group; 2020;10:7648. 10.1038/s41598-020-64523-1

31. Cheng C, Zhao H, Tian W, Hu C, Zhao H. Predicting the expression level of Ki-67 in breast cancer using multi-modal ultrasound parameters. BMC Medical Imaging. 2021;21:150. 10.1186/s12880-021-00684-3

32. Luporsi E, André F, Spyratos F, Martin P-M, Jacquemier J, Penault-Llorca F, et al. Ki-67: level of evidence and methodological considerations for its role in the clinical management of breast cancer: analytical and critical review. Breast Cancer Res Treat. 2012;132:895–915. 10.1007/s10549-011-1837-z

